# The successful introduction of a hand dermatitis clinic to reduce occupational dermatoses during the covid-19 pandemic

**DOI:** 10.1101/2021.02.04.21250767

**Authors:** Sarah Walsh, Siobhan Carey

## Abstract

**Background:** The SARS-Co-V pandemic has necessitated strict hand hygiene practices in healthcare settings. An increased incidence of staff presentations of occupational dermatitis in our institutions prompted the establishment of a drop-in access daily hand dermatitis clinic.

**Objective:** to document the incidence and severity of cases of occupational dermatitis arising in healthcare workers during the first wave of the pandemic, and to reduce the impact of this with early assessment and treatment.

**Methods:** an open-access daily staff clinic run by dermatologists was established, and demographic and clinical data collected from each clinical encounter.

**Results:** 532 staff attended the clinic over a 6 week period. This compared to 7 staff presenting to occupational health over the same period in 2019. The majority were females 81%,and ward based 51%. ITU and A&E staff represented 15%. The prevalence of occupational hand dermatitis was 88%. 52% were classed as mild, 26% as moderate 26% and 9% severe/very severe.

**Conclusions:** the pandemic and associated hand hygiene practices have led to a substantial increase in presentations of occupational hand dermatitis. A drop-in clinic proved an effective way of allowing staff to access treatment in a timely fashion.

## Introduction

The importance of hand hygiene in the reduction of viral transmission has been recognized with one large meta-analysis suggesting optimum hand hygiene can lead to a 24 % reduction in viral transmission (1). The physical action of hand washing (for minimum 20 seconds) disrupts the lipid barrier of the covid-19 virus and thus hand hygiene is one of the most important preventative measures to reduce and ultimately stop the spread of covid-Frequency of hand-washing has increased, particularly in healthcare settings. Prior to the covid-19 pandemic, one study suggested that the estimated prevalence of occupational dermatoses was 20% for clinical and 7% for non-clinical staff in a population of UK health care workers (2). An increased prevalence of hand dermatitis in those employed in this sector has also been observed (3). A study from China has demonstrated that risk of hand dermatitis rises with increasing numbers of episodes of hand-washing performed per day, suggesting a “dose-response” effect (4, 5).

Studies of occupational hand dermatitis during the pandemic have largely been survey-based, and performed in a post-hoc fashion. At the outset of the pandemic in our institution, we established an in-person “teach and treat” hand dermatitis (HD) clinic for all staff, with two objectives: firstly, to treat occupational hand dermatitis in a timely fashion, convenient to staff, enabling them to remain at work; secondly, to provide education regarding good hand care which would be propagated throughout the organization by clinic attendees.

## Methods

In conjunction with the Occupational Health department, a simple proforma was developed to record the staff member’s age, location of work, role in the hospital, and history of eczema. Brief targeted education regarding the importance of regular application of emollient, and more intensive overnight treatment was designed, supplemented by a written information sheet for the staff member to take away.

Two dermatologists staffed the clinic for one hour, initially daily, reducing to 3 times a week as demand reduced, in an appropriately-sized clinical area within the hospital. Personal protective equipment was worn, and social distancing measures adhered to. The proforma was completed by the attendees, followed by examination of the hands by the attending clinician. A Physician Global Assessment score was assigned, according to disease severity, followed by delivery of the brief educational intervention. Topical steroid of appropriate potency was prescribed if required, with instructions given as to correct dose and use.

## Results

532 staff members attended the HD clinic during a six-week period from 26th March-6th May 2020. In the same period in 2019, 7 patients had presented to occupational health with occupational hand dermatitis. In our cohort, the majority were females 81% (432), with a median age of 42.5 years. The majority of staff were ward based 51% (272). ITU and A&E staff represented 15% (82), theatre staff 5% (28) and the remainder of staff were outpatient, office based or other (laboratory, stores etc.).

Nursing staff represented 33% (177) of attendees, doctors 22% (104), allied health professionals such as physiotherapists and dieticians 21% (112). The remainder were classified as other (porters, administrative staff, scientists and housekeeping). 33% (178) of attendees reported a previous history of eczema.

The prevalence of hand dermatitis was 88% (468) which was graded as mild 52% (276), moderate 26% (139) and severe/very severe 9% (50). Treatment with topical steroid was required in 42% (225) of staff. Of the 225 requiring topical steroids, the majority required moderate 49 %(111) potency steroid. 44% (100) required potent steroid, highly potent steroid 1.8 %(4) and 0.8 %(2) required a mild potency steroid.

A small subset of attendees were approached to evaluate the response to the HD clinic. All (10/10) were very satisfied with the service. Copious informal feedback from colleagues, including those in Occupational Health, suggests that this intervention was highly valued by colleagues during the pandemic.

## Conclusion

Review of the literature indicates that this cohort represents the largest reported number of healthcare workers evaluated in person for hand dermatitis during the pandemic. Compared to previous years, the incidence of occupational hand dermatitis increased significantly. This affected all staff groups and in almost half required treatment with topical steroid. We hope now to evaluate whether that this preemptive approach to addressing occupational hand dermatitis during the pandemic resulted in a lower disease burden, and increased staff retention at work.

## Data Availability

All data reported in this manuscript is available by application to the authors.

